# Remote Real Time Digital Monitoring fills a Critical Gap in the Management of Parkinson’s disease

**DOI:** 10.1101/2024.12.12.24318893

**Authors:** Aarushi S. Negi, Shreesh Karjagi, Laura Parisi, Kerry W. Daley, Annie K. Abay, Aryaman S. Gala, Kevin B. Wilkins, Shannon L. Hoffman, Margaret S. Ferris, Hengameh Zahed, Gaurav Chattree, Bianca Palushaj, Helen M. Bronte-Stewart

**Affiliations:** Department of Neurology and Neurological Sciences, Stanford University School of Medicine, Stanford, CA, United States; Department of Neurosurgery, Stanford University School of Medicine, Stanford, CA, United States

**Author notes:** Corresponding Author: Helen Bronte-Stewart, Department of Neurology and Neurological Sciences, Center for Academic Medicine, Rm 254A, 453 Quarry Rd, Stanford University School of Medicine, Stanford, CA 94305. These authors contributed equally to this work (Aarushi S. Negi, Shreesh Karjagi).

## Abstract

**Background and Objectives:** People with Parkinson’s disease (PWP) face significant gaps in care. Limited neurologist access, infrequent clinic visits, and inadequate symptom measurement culminate in suboptimal therapy and high morbidity. Quantitative Digitography (QDG) addresses these care gaps by providing validated, quantitative metrics of all motor signs in Parkinson’s disease (PD) in real-time through a 30-second repetitive alternating finger tapping (RAFT) task. We investigated the feasibility and clinical relevance of remote QDG monitoring in individuals with movement disorders.

**Methods:** In this prospective cohort study, participants with suspected or clinically established PD were referred by neurologists from Stanford Movement Disorders Clinic, with one participant recruited outside Stanford. Participants were asked to complete at-home QDG-RAFT for 30 days and were administered usability and quality-of-life questionnaires throughout testing; descriptive statistics summarized compliance and questionnaire responses, and a Spearman correlation assessed the relationship between QDG Mobility Score and MDS-UPDRS Part II scores. The primary outcome was compliance with once-daily testing for at least 16/30 days of remote monitoring.

**Results:** 30 participants (24 with clinically diagnosed and 6 with suspected PD at time of referral) were included. 100% of participants demonstrated compliance with once-daily testing for at least 16/30 days. Adherence rates for once-daily and twice-daily testing were 96.2% and 82.2%, respectively, with 96% of participants rating once-daily testing as easy. The QDG Mobility Score correlated strongly with patients’ self-reported impact of motor symptoms on Activities of Daily Living (ADLs) per the MDS-UPDRS II (ρ=-0.61 [−0.88, −0.16], p=0.004). QDG-RAFT documented motor asymmetry and impairment from 1-month pre-diagnosis to 20-years duration of PD. The QDG Mobility Score reflected motor improvement and deterioration after adding or removing respectively a single tablet of carbidopa/levodopa.

**Discussion:** Participants showed excellent compliance with remote QDG monitoring and found the system easy to use. The QDG Mobility Score was highly correlated with ADLs, captured motor complexities across a broad disease duration range, and detected responses to minor therapy adjustments. A pivotal advancement in PD care, QDG offers providers an accessible, comprehensive tool to remotely monitor motor symptoms, optimize treatment regimens, and bridge care gaps created by infrequent clinic visits and subjective symptom assessment.

## Introduction

Parkinson’s disease (PD) presents a complex healthcare challenge. Among the >1.2 million Americans living with PD,^1^ 40% lack access to a neurologist.^2–4^ Three fundamental limitations undermine current care: infrequent, brief in-person clinic visits, a complex clinical examination, and a lack of widely available remote monitoring systems. People with PD (PWP) are usually evaluated in person every three to six months.^5^ As PD is a progressive disease, the treatment plan set by the neurologist at one visit may become subtherapeutic by the next, which leaves PWP to adjust their medications themselves; a practice that results in unstable dopamine levels and swings from under- to over-treatment. This results in an increased incidence of falls, fractures, and neuropsychiatric complications such as confusion and hallucinations, the complications of which can lead to death.^6–8^. Furthermore, the Movement Disorder Society-Unified Parkinson’s Disease Rating Scale (MDS-UPDRS)^9^ motor examination (Part III) is comprehensive but subjective and variable within and among raters.^10^ Most health care providers are neither trained nor have time to administer it during short visits, and PWP lack objective, comprehensive motor monitoring in between.^11^ The reliance on subjective recall rather than objective data about symptoms between visits can also lead to suboptimal medication management and treatment adjustments.^12^ There is a need for remote, objective monitoring technologies that fill the majority of the PWP journey, which occurs outside of clinical visits.^13^ This will provide a more comprehensive approach to symptom tracking and more frequent adjustments of therapy, which will facilitate optimal disease management.^14^

Quantitative Digitography (QDG) solves this critical unmet need by providing validated, quantitative metrics of all motor signs in PD, remotely and in real-time.^15–19^ From a brief 30-second repetitive alternating finger tapping (RAFT) task, QDG delivers high-resolution motor metrics that correlate with the MDS-UPDRS III scores and sub-scores, track symptom progression, and demonstrate sensitivity to adjustments in therapy.^16,17,19,20^ The integrated QDG system improves the assessment and management of PD by enabling point of care remote monitoring and results available in the electronic health record (EHR) in real time,^21^ allowing clinicians flexibility in testing schedules and to optimize PWP-specific treatment plans in between in-person visits. The QDG system has been granted Breakthrough Device Designation by the United States Food and Drug Administration (FDA).

In this prospective cohort study, we demonstrate the feasibility and clinical relevance of remote digital monitoring using the QDG system in people with movement disorders, when referred by neurologists. The primary outcome was compliance with performing one test per day for at least 16/30 days of remote monitoring. Secondary outcomes included adherence with testing once and/or twice per day, user experience, correlation between the QDG Mobility Score and the participant’s perception of the impact of their motor function on Activities of Daily Living (ADLs), and QDG’s sensitivity to detect motor changes after small adjustments in therapy.

## Methods

### Participants

Inclusion criteria: people with suspected or clinically established PD, who were over 18 years of age, able to follow instructions, and provide informed consent. Exclusion criteria: people unable to perform the task due to pain and/or musculoskeletal injury or disease.

### Standard Protocol Approvals, Registrations, and Patient Consents

This study was approved by the Stanford University Institutional Review Board (IRB) in accordance with recognized ethical guidelines (IRB eProtocol #60883). All participants provided written informed consent prior to enrollment in the study. The study was not registered as a clinical trial, as it does not involve the testing of a health-related intervention within the scope of a regulated trial registry.

### QDG System

The QDG system consists of a Bluetooth-enabled digitography device (KeyDuo), a patient-facing mobile application, a HIPAA-compliant cloud web service and customized algorithm (PRECISE), and an EHR-integrated web dashboard.^21^ The KeyDuo comprises adjacent tensioned, engineered levers, which can sense the displacement and timing of lever motion with a sampling rate of 201 Hz and accuracy of 0.12 mm throughout the device’s range of motion. Patients interface with the QDG mobile application (operating system iOS 16.4-current) to initiate a test, enter therapy settings, and complete the QDG-RAFT task. The data are transferred from the KeyDuo to the QDG mobile application using Bluetooth. The mobile application conducts a device calibration-specific raw data transformation, checks for errors, and collates medication and deep brain stimulation (DBS) settings. The data are stored in the HIPAA-compliant cloud service, where each RAFT test is queued for analysis by the QDG PRECISE algorithm. The QDG at-home apparatus included a KeyDuo, iPad mini, palm rest, and cable (to power the KeyDuo via iPad). The QDG mobile application was installed on the iPad in advance.

#### QDG Algorithm and Motor Data

The PRECISE algorithm analyzes KeyDuo raw data to extract press and release amplitudes and speeds, their coefficients of variation (CV = standard deviation/mean), inter-strike intervals (ISI) and ISI CV, as well as release and dwell times (durations at the top of the release and base of the press phases, respectively).^17–21^ Sub-algorithms detect strikes generated by rest or action tremor, analyze the duration, average amplitude, and frequency of tremor, and remove those strikes from the analysis of voluntary movements.^22^

The measures from voluntary strikes yield quantitative, validated metrics of bradykinesia, rigidity, and gait impairment.^18,19^ QDG provides four validated metrics of bradykinesia: tapping frequency, press amplitude, press speed, and press amplitude variability (press amplitude Coefficient of Variability (CV)), which captures the deterioration of press amplitude over time, known as the sequence effect. The validated QDG rigidity metric is the release speed^18^, and the metric for gait impairment and freezing behavior is arrhythmicity.^19^ The algorithm also quantifies the number and duration of freezes, termed freezing of RAFT (FoRAFT).

Metrics are averaged across the 30-second trial for each finger, averaged between fingers for each hand, and used to calculate the QDG Mobility Score. The QDG Mobility Score (ranging from 0 – 100) is a statistically derived composite score that represents overall motor performance by statistically weighting QDG voluntary movement metrics and normalizing them against age-matched healthy controls; a higher score represents better performance. A separate Tremor Severity Score (ranging from 0 – 100) is calculated based on percent duration and average amplitude of tremor strikes during the task; a higher score represents greater tremor severity.^21^

#### Clinician Dashboard

QDG output metrics are transmitted back to the web service and are displayed through an interactive SMART-on-FHIR Dashboard, a protocol for interoperability across EHR systems. The dashboard displays single test results (Supplementary Figure S1), medication schedule, and metric data over any time range. The dashboard was available on the web portal and was embedded in the Electronic Health Record (EHR), allowing health care providers to access QDG data within the PWP’s chart in real-time.

### Experimental Protocol

Figure 1 outlines the study protocol and flow. Participants were screened for the study inclusion and exclusion criteria over the phone. For information on questionnaires administered in the study, see Supplementary Materials.

**Figure 1.**
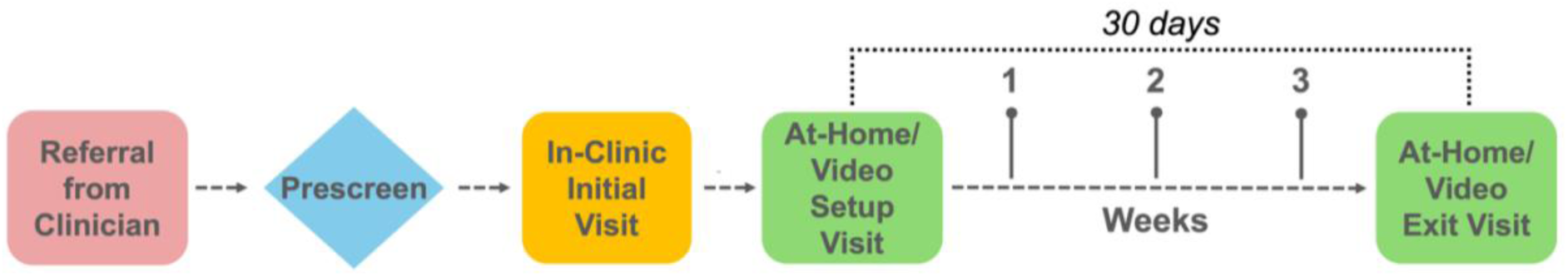
QDG Remote At-Home Study Protocol. During the visits, the MDS-UPDRS II and questionnaires on QDG system usability, design feedback, and PD symptom tracking were administered (Supplementary Table 1).

#### Baseline Clinic Visit

At the initial clinic visit, participants received an overview of the study purpose and protocol, including participant responsibilities, and provided written informed consent. Participants performed a baseline QDG-RAFT task and were assessed with the MDS-UPDRS III by a certified rater.

#### At-home Setup Visit

Research team members initially traveled to participants’ homes to assist with setup and training on use of the QDG system. As the study progressed, setup and training expanded to include a remote option via video conferencing. The research team first reviewed the QDG user guide with the participant and trained them on mobile app navigation. Participants were then asked to set up and execute the QDG-RAFT task on their own to ensure proper use of the system. Once a participant-initiated QDG-RAFT test had been successfully completed, research personnel confirmed the participants’ testing schedule based on the recommendation of the referring clinician. PWP on medication were instructed to complete two tests per day, once in their worst “off state” and once in their best “on state,” whereas participants who did not take medication were asked to test once a day. The participants then reported their baseline “motor experiences of daily living” using the MDS-UPDRS II (ADL) scale (Supplementary Table 1). The 30-day remote testing period commenced after this visit.

#### Remote monitoring Phase

Routine check-ins were conducted either in-home or via video conferencing once per week for 4 weeks with research team members (Figure 1). At each check-in, the participant completed questionnaires regarding potential adverse events (Custom Adverse Event Questionnaire), testing adherence and usability (In-Home Usability Testing and User Feedback Questionnaire), and the MDS-UPDRS II (Supplementary Table 1). Additionally, research personnel observed participants’ execution of the QDG-RAFT task to confirm that they maintained correct task performance and retrained participants as needed.

#### Exit Visit

Upon completion of the remote monitoring period, research team members visited the participant’s home to complete their final check-in, which included routine weekly check-in items as well as a Symptom Tracking and Communication Survey (Supplementary Figure S4), PD History Questionnaire, Design Feedback Questionnaire, and an Exit Interview (Supplementary Figure S3). Further along in the study, the exit visit could be completed over video conferencing if the participant lived further away.

### Statistical Analysis

Statistical analyses were performed using Python (v3.12.5) with SciPy (v1.11.3) and NumPy (v2.0.0) libraries. Spearman rank correlation was used to evaluate the relationship between QDG Mobility Scores and MDS-UPDRS Part II scores. A bootstrapping approach with 10,000 resamples was used to estimate the 95% confidence interval (CI) for the Spearman correlation coefficient. A p value < 0.05 was considered significant. No correction for multiple comparisons was required since only one statistical test was run.

### Data Availability Statement

The datasets used and/or analyzed during the current study may be shared (anonymized) from the corresponding author on reasonable request for research studies with a defined scientific question and plan pertaining to the use of the data.

## Results

### Participant Demographics

Thirty participants (23 males) provided informed consent and entered the study; twenty-nine were referred by neurologists at Stanford Movement Disorders Clinic and one was recruited from outside of Stanford (Table 1). One participant was lost to follow-up, and four participants partially completed or exited the study early due to technical difficulties or schedule conflicts (Supplementary Figure S5). Twenty-five participants completed the full 30-day protocol and were included in the final analysis. Participants were referred for multiple clinical purposes, ranging from medication response monitoring to pre-diagnostic assessment. The cohort also captured a broad range of Parkinson’s disease duration, from people undiagnosed but with suspected Parkinsonism to those with clinically established PD of long duration and with motor fluctuations. The cohort exhibited well-controlled symptoms on therapy with medications and/or DBS (Table 1).

**Table 1.** Participant Demographics and Clinical Characteristics.

| Characteristic | n (%) |
| --- | --- |
| **Age** |  |
| Mean ± SD (years)* | 67.0 ± 8.9 |
| Range | 50–83 |
| **Sex** | N = 30 |
| Male | 23 (76.7) |
| Female | 7 (23.3) |
| **Race/Ethnicity** | N = 30 |
| White | 24 (80.0) |
| Asian | 5 (16.7) |
| Hispanic or Latino | 1 (3.3) |
| **Diagnosis at Time of Referral** | N = 30 |
| Established PD | 24 (80.0) |
| Essential Tremor | 2 (6.7) |
| Tremor† | 2 (6.7) |
| Mild Cognitive Impairment | 1 (3.3) |
| Dream Enactment | 1 (3.3) |
| **Years Since PD Diagnosis\*** | N = 25 |
| < 5 years | 7 (28.0) |
| 5-10 years | 4 (16.0) |
| 10-15 years | 10 (40.0) |
| > 15 years | 4 (16.0) |
| **ON Therapy MDS-UPDRS III§** | N = 28 |
| Mean ± SD | 15.3 ± 7.1 |
| Range | 2–26 |
| **Reason For Referral** | N = 30 |
| Medication response monitoring | 18 (60.0) |
| Motor fluctuation | 12 |
| Adherence | 4 |
| Medication initiation | 2 |
| Motor monitoring | 7 (23.3) |
| Pre-diagnostic assessment | 4 (13.3) |
| DBS programming | 1 (3.3) |

\*Calculated for the PD cohort at the time of In-Clinic Visit 1.
†One participant presenting tremor at time of referral received a PD diagnosis during the study.
§One MDS-UPRS III assessment performed OFF therapy.

### QDG Mobile Application, Task Setup, and Execution

Figure 2 represents the participant interface with the QDG remote system. Participants set up a QDG account and inputted their anti-parkinsonian medication schedule and DBS model and settings (if applicable) (Figure 2A). They were then trained on setting up the QDG system (Figure 2B). This included finding an optimal location where they could perform the task in a seated position with their forearm flexed at roughly 90 degrees; the device was placed on a flat, stable surface with the depressions on the levers aligned with the fingertips when the wrist was supported, such that the wrist was in a neutral position. They established a Bluetooth connection between the KeyDuo and QDG Mobile App, initiated a test, and confirmed whether they were on or off medication and/or DBS when applicable. Execution of the task was cued by an instructions screen, which prompted participants to position themselves for their first test (Figure 2C). Participants were told to fully press and release their index and middle fingers in an alternating fashion, tapping as fast and as consistently as possible. Auditory “Go” and “Stop” cues were given through the app to start and stop RAFT, respectively. Testing always started with the right hand followed by the left (Figure 2C). Upon saving both tests and returning to the mobile app home screen, data was automatically sent to a secure cloud service for analysis.

**Figure 2.**
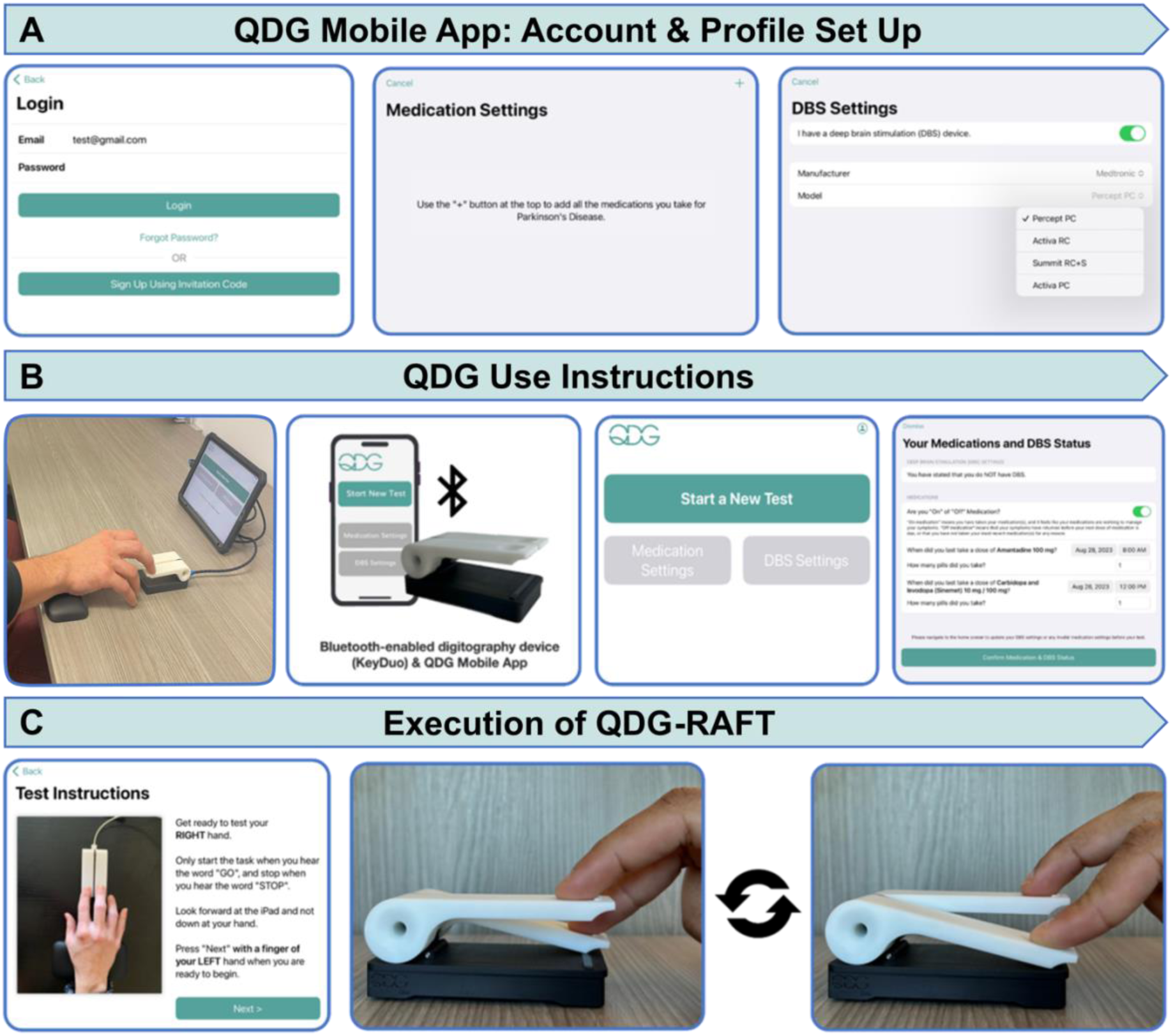
QDG Mobile Application Use and Task Execution. A) Workflow for mobile application account set-up. B) QDG task initiation and setup instructions. C) QDG-RAFT task execution.\

All participants were able to set up the system correctly at the first week check in; this included hardware connection, therapy screen navigation, and task initiation. All participants achieved complete task proficiency by the second week, despite occasional initial challenges with Bluetooth connectivity (Supplementary Figure S2).

### Compliance, Adherence, and User Experience

Participants demonstrated excellent compliance with remote QDG testing, with 100% of participants completing at least one test a day for 16/30 days (Figure 3A). Participants maintained adherence rates of 96.2% (N=25) for completion of one test per day and 82.2% (N=24) for two tests per day (Figure 3B). Only PD participants on dopaminergic medication were asked to perform two tests per day.

**Figure 3.**
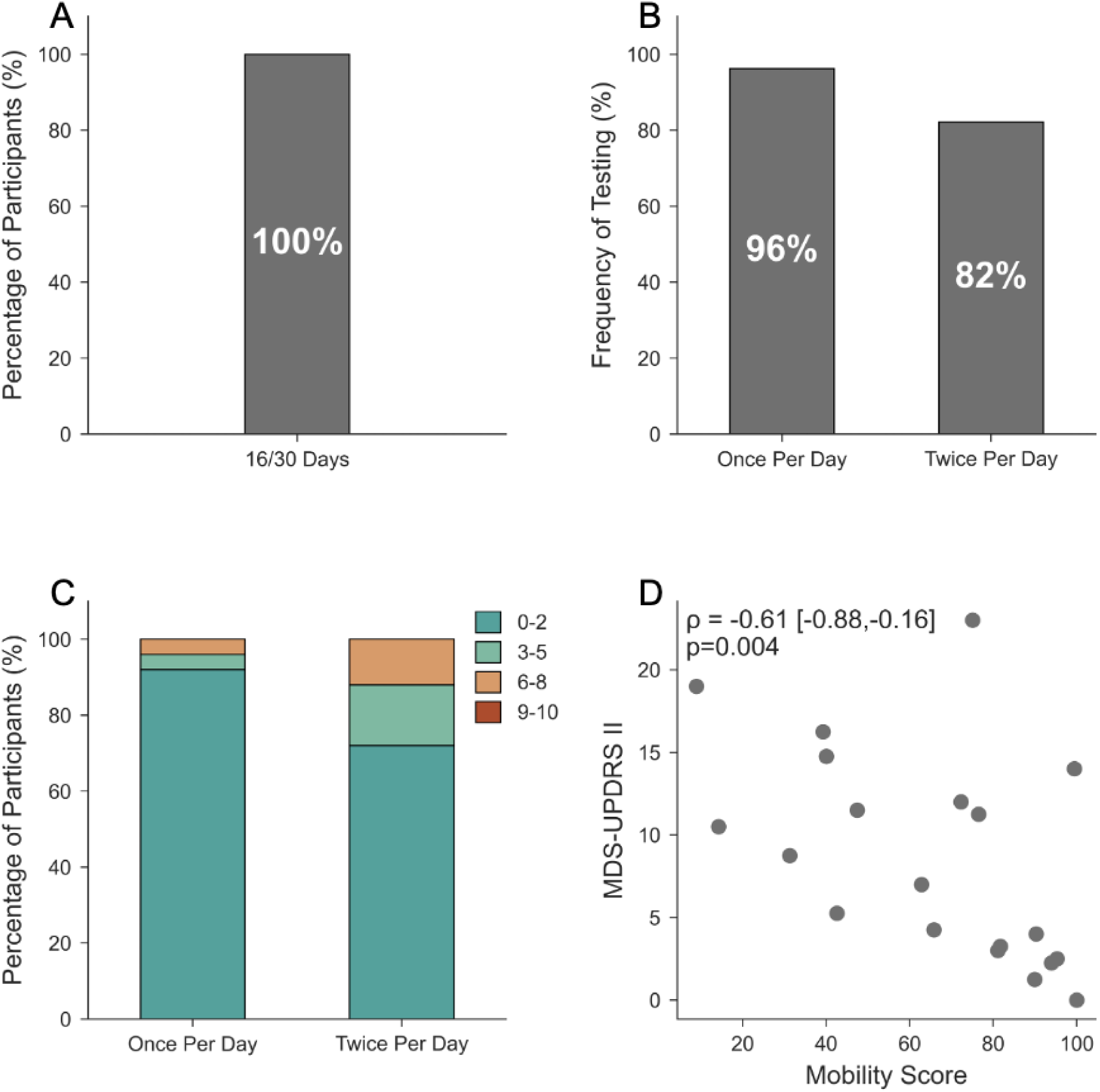
(A) Compliance with a test performed on at least 16/30 days; (B) Adherence to testing once-daily (N=25) and twice-daily (N=24), (C) User reported difficulty ratings with performing QDG-RAFT (0=extremely easy, 10=extremely difficult). For visualization purposes, responses were categorized as Extremely Easy (0-2), Moderately Easy (3-5), Moderately Difficult (6-8), or Extremely Difficult (9-10); (D), QDG Mobility Scores correlated with MDS-UPDRS Part II scores (ρ= −0.61 [−0.88, −0.16], p=0.004, n=20).

User experience assessments revealed that all but one of the participants (24/25, 96%) rated the ease or difficulty of doing one test per day as easy (<5/10 on the Likert scale, (Supplementary Figure S3); the other participant rated it with a 6/10. For twice per day testing, 88.0% (22/25) rated it as <5, and 12.0% (3/25) rated it between 6-8. None of the participants rated either testing schedule greater than 8 (Figure 2C).

Participants reported various strategies to facilitate scheduled testing. These included setting alarms, incorporating tests into written schedules, and aligning testing with medication schedules. Participants who found twice per day testing moderately difficult primarily reported travel schedules and Bluetooth connectivity issues as barriers to full adherence.

### QDG Mobility Score correlated with MDS-UPDRS II (ADL) score

In the PD cohort, the QDG Mobility Score (averaged across hands, and across weeks) demonstrated a high, significant correlation with participants’ MDS-UPDRS II, averaged over four weeks; ρ=-0.61, 95% CI: [−0.88, −0.16], p=0.004, N=20, Figure 2D. Higher QDG Mobility Scores reflected better abilities in daily living tasks (lower MDS-UPDRS II scores, Figure 2D).

### QDG reflects time course of PD

There was a wide range of disease duration among the participants with diagnosed or suspected PD, who were referred for participation (pre-diagnosis to 20 years, Table 1). Figure 4 demonstrates representative QDG traces from the more and less affected sides from five participants with different durations of disease.

**Figure 4.**
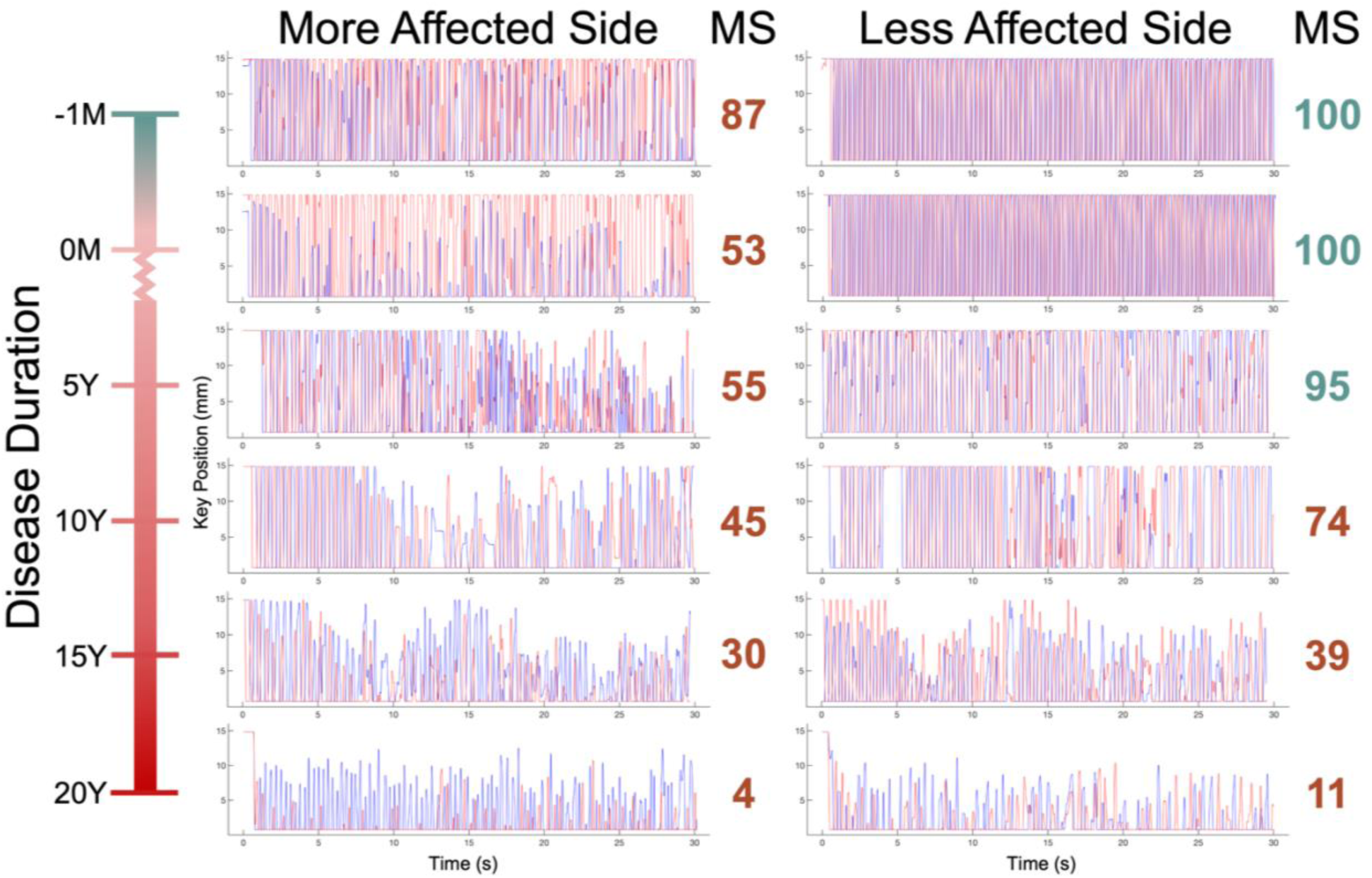
QDG-RAFT traces and Mobility Scores (MS) from the more and less affected sides of representative participants with different durations of PD (−1 month, 0 months, 5, 10, 15, and 20 years). Each RAFT trace displays the amplitude of lever strikes (millimeters) over time (seconds); the blue and red strikes represent the index and middle fingers on right hand traces (vice versa on left hand). A normal MS (>92) is in green and abnormal in red. The –1- and 0-month traces are off therapy, while 5- to 20-year traces are on therapy.

The top two traces are from the same participant, who was referred for remote monitoring on the day of initial presentation of intermittent left-hand tremor, only when walking. The initial QDG-RAFT test revealed an abnormal Mobility Score (MS) of 87 (normal > 92) on the left (more affected, MA) hand, and normal MS (100) on the right (less affected, LA) hand. One month later, on the day of their PD clinical diagnosis, the same participant’s performance of the MA hand was worse (MS = 53), demonstrating progression of disease in the month prior to the clinical diagnosis, whereas the performance remained normal in the LA hand (MS = 100). In addition to asymmetry, there was evidence of differing performances between the two fingers (blue and red strikes) on the MA hand at the 2^nd^ test (time of diagnosis) of this participant, which we have labelled “finger dissociation.” QDG traces from a representative participant with PD for 5 years, on therapy, showed asymmetry of performance between hands; the MS was 55 on the MA and 95 on the LA hands respectively. There was evidence of the sequence effect in both fingers only on the MA side after 17 seconds of RAFT. After 10 years of PD, a representative trace (on therapy) demonstrated abnormal performance on both sides, although still asymmetric (MS = 45 | 74). The sequence effect occurred on the MA side after only 7 seconds of RAFT, and led to a freezing event (FoRAFT) at 12 seconds shown by the pause in movement of the red finger strike at the bottom of the lever press. After this, the performance was more irregular in amplitude and frequency. FoRAFT and the sequence effect were also evident on the LA side. For the representative participant with a 15-year disease duration, on therapy, the performance was worse bilaterally with less difference between sides (MS = 30 | 39). Finger dissociation and the sequence effect were evident bilaterally and occurred early, and there was lower amplitude tapping in general. Lastly, the representative participant tested after 20 years of PD demonstrated the lowest MS (MS = 4 | 11), low amplitude RAFT bilaterally with prominent finger dissociation, irregular amplitude and frequency, and very early reduction in amplitude that resulted in FoRAFT bilaterally (red fingers).

### QDG tracks small, patient-initiated adjustments of medication

Figure 5 demonstrates the change in the QDG-RAFT MS after the addition (Figure 5A) or reduction (Figure 5B) of one tablet of immediate release carbidopa/levodopa (CD/LD 25/100) during remote monitoring.

**Figure 5.**
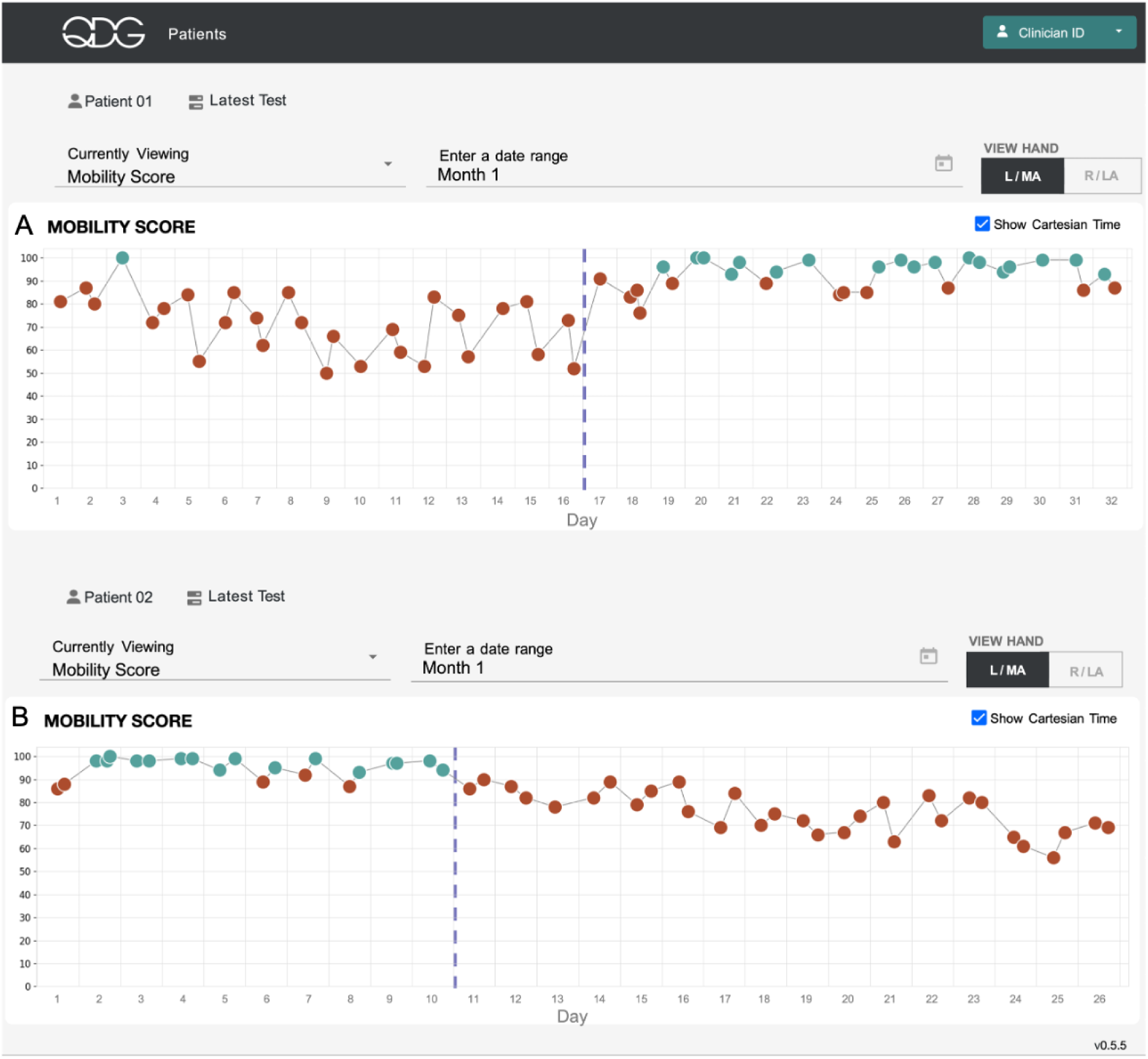
QDG dashboard showing Mobility Scores. (A) Participant 1 [adapted from ^21^] and (B) Participant 2’s more affected left-hand scores. Green circles indicate normal values, red circles abnormal values. Dashed lines mark therapy adjustments.

The MS on the More Affected side of a participant with early-stage PD improved from abnormal (71.9 ± 13.2) to normal (92.6 ± 6.5) after the addition of one tablet of CD/LD 25/100mg on day 17 to a regimen of three doses a day plus Sinemet-CR at night (Fig. 5A, dashed line). The improvement was evident on the same day and the day-to-day variability also decreased. In contrast, Fig. 5B demonstrates a decrease of the MS after a participant decided to stop medication altogether after the first 10 days of monitoring due to involuntary movements. The participant had advanced PD, treated with DBS (14 years post-diagnosis) and minimal medication (0.5 tablet CD/LD 25/100mg twice a day). Their average MS of 95.0 ± 4.6 deteriorated after the medication withdrawal (77.0 ± 10.8). The progressive decrease of the MS over 3 weeks may reflect the long duration levodopa response.^23^

### Symptom tracking and communication survey

Participants who had clinically established PD at the time of referral answered a survey focused on symptom tracking and communication at the exit interview (Supplementary Figure S4). 84.2% (16/19) of participants responded that prior to the study, they did not have a robust monitoring method for their symptoms, and either did not track their symptoms at all, or tracked them mentally. One participant recorded symptoms with a diary, and one tracked tremor using the Apple watch. However, the majority (57.9%, 11/19) wished they had a more objective way to communicate their symptoms or change in symptoms with their clinician. With respect to standard of care, 52.6% (10/19) of participants were assessed by their neurologist using the MDS-UPDRS III no more than twice annually, with 15.8% (3/19) of participants seeing their neurologist no more than once per year.

## Discussion

Remote monitoring using the QDG system demonstrated 100% compliance with the requirement to perform one test a day for at least 16 out of 30 days. There were strong test adherence rates of 96.2% for once-daily and 82.2% for twice-daily testing over the 30-day period. All participants were able to set up and complete the task correctly at their first weekly check-in, and 96% of participants rated once-daily testing as easy. Referrals came from a Movement Disorders neurology clinic, based on individual neurologist’s prescription and need for adjunctive information to their clinical assessments for a range of movement disorders diagnoses and monitoring objectives. Among the participants with PD, QDG reflected symptom severity and asymmetry throughout a broad range of disease durations, from pre-diagnosis through 20 years post-diagnosis. The QDG Mobility Score was highly correlated with the PWP’s reported ADL impairment, and QDG’s high resolution metrics were sensitive to small medication adjustments.

The clinical impact of QDG monitoring was highlighted through several key findings. First, the strong correlation between the QDG Mobility Score and MDS-UPDRS II validates its real-world relevance. The Mobility Score provides a sensitive quantification of motor symptoms that also meaningfully reflects a patient’s functional status and the impact of those symptoms on everyday activities. Next, QDG measured and conveyed the motor complexities of PD over a broad range of disease duration. Pre-diagnosis, QDG captured high asymmetry between the MA and LA hands, a valuable insight that could aid clinicians in diagnosis, and documented progression of disease severity even before diagnosis, which would not have been noticed or documented without remote monitoring in between visits. It is well documented that the sequence effect in gait (progressive shortening of stride length) leads to Freezing of Gait (FOG) and Freezing of Upper Limb movement (FOUL).^15,24–27^ In this study QDG-RAFT demonstrated that the sequence effect also led to FoRAFT; it was more severe and occurred earlier in the task at longer durations of disease.

Daily monitoring with QDG-RAFT revealed critical therapeutic windows typically missed in routine care – from quantifying immediate motor improvement after a single tablet of CL/LD in early-stage PD, to capturing the gradual decline when a DBS participant abruptly stopped a single tablet of CD/LD. To our knowledge, QDG represents the only quantitative point of care remote monitoring system to capture the motor effect of small adjustments in CD/LD dosing in early and advanced PD, while achieving the high compliance needed for routine clinical care. QDG’s ability to detect changes in dopa-responsive motor symptoms positions it to play a similar role in PD care as continuous glucose monitoring (CGM) does for diabetes management. In both diseases, patients must adhere to strict, time-sensitive medication schedules to maintain glucose or dopamine levels respectively within defined physiological ranges. While continuous glucose monitoring (CGM) has advanced diabetes care by providing real-time feedback for insulin dosing,^28^ a similar tool has been so far lacking in PD care. We believe that QDG comprehensive motor monitoring will ameliorate PD care by giving the provider and PWP actionable insight into their motor function, enabling healthcare providers to optimize therapy in real-time across the disease spectrum.

QDG addresses a major need for people with movement disorders: the clinician-patient gap in assessment and communication. In our PD cohort, 52.6% of participants were assessed by their neurologist using the MDS-UPDRS III no more than twice annually, and 15.8% of participants saw their neurologist no more than once per year. Although the majority of participants did not monitor their disease symptoms prior to the study, most expressed a desire for more objective ways to monitor and communicate symptoms to their healthcare providers. QDG can only provide clinically meaningful value if patients and providers adopt it. The Centers for Medicare and Medicaid Services (CMS) requires patients to obtain a physiological parameter (i.e., blood pressure, glucose) on at least 16/30 days for providers to gain additional revenue through remote physiological/therapeutic monitoring (RP/TM) codes.^29^ QDG met this CMS requirement with 100% of participants testing at least 16/30 days, further strengthening its viability for widespread clinical adoption.

To our knowledge, QDG is the first comprehensive, real-time, remote monitoring system for people with movement disorders, offering validated, high-resolution metrics of motor function and flexible use case scenarios for neurologists. Current wearable systems for passive monitoring could be a complementary solution to QDG monitoring. However, these face some limitations that may impact their clinical utility on a daily basis. None can capture all motor symptoms of PD with one device and in real-time. Users are instructed to wear the sensor for at least six hours per day and over the course of six days to produce reliable data.^38,39^ The averaged retrospective data is reported asynchronously to the provider and not through the EHR. Most monitor a single wrist’s movement, and this may not reflect or capture movements involving multiple body parts. Wearable systems that require multiple simultaneous sensors can create usability barriers due to the complexity and inconvenience. ^30 36,37^

Finger tapping performance has been shown to reflect overall PD severity and is an early or prodromal disease marker.^16,19,31–35^ QDG-RAFT provides crucial insight into bilateral characteristics of PD, such as asymmetry, which would be missed by single-point sensors. The QDG system’s direct integration with the electronic health record allows clinicians to access QDG data synchronously with their management of each patient, thus not impacting their over-capacity workflow. QDG’s comprehensive nature further expands the scope of its accessibility amongst physicians, allowing general neurologists and primary care physicians to rapidly glean clinical insights that would typically require examination by a movement disorders specialist.

### Limitations

This initial remote monitoring trial investigated the feasibility of daily remote monitoring using QDG and included a relatively small sample size. A larger study may expand QDG compliance and usability. Another limitation early in the study was the need for research personnel to travel to participants’ homes for QDG system setup, training, and exiting. Once this was determined to be successful, the study expanded to recruit out-of-state participants, utilize remote video conferencing, and leverage device shipment to and from participants’ homes. This enabled geographical expansion of remote monitoring using the QDG system.

## Conclusion

QDG is a comprehensive, real-time, remote monitoring system for people with neurological disorders that offers a flexible set of use case scenarios for neurologists and other health care providers. All study participants successfully completed at least 16/30 days of testing and demonstrated high rates of adherence to both once-daily and twice-daily testing schedules for 30 days. Participants were able to effortlessly interact with the system, and 96% rated once-daily testing as easy. In addition, the QDG system generated high-resolution data that closely correlated with PWP’s self-reported ADLs and differentiated motor impairment and asymmetry from those pre-diagnosis to others up to 20 years post-diagnosis. A majority of participants expressed the desire to have a more objective way to communicate their symptoms, or change in symptoms, with their provider. The QDG system represents a critical advancement in PD care, equipping providers with a portable, accessible tool to monitor the comprehensive set of motor symptoms remotely, optimize therapeutic regimens, and bridge the care gaps created by infrequent clinic visits and subjective symptom assessment. By integrating seamlessly into clinical workflows without adding complexity, QDG redefines the standard of care for movement disorders. Its unique ability to scale across disease stages and sense small adjustments in therapy positions QDG as an indispensable resource in the ongoing effort to improve the management of PD.

## Acknowledgements

The authors would like to thank all research participants who generously contributed their time to this study. We would also like to thank Simon Revlock and Christopher Coriz for their contribution to QDG hardware design; Paul Schmiedmayer, Vishnu Ravi, and Warren Hiemstra for their software development work; and Morgan Hunt for project management.

