## Supplementary Materials for "Remote Real Time Digital Monitoring fills a Critical Gap in the Management of Parkinson’s disease"

### Methods

### QDG System

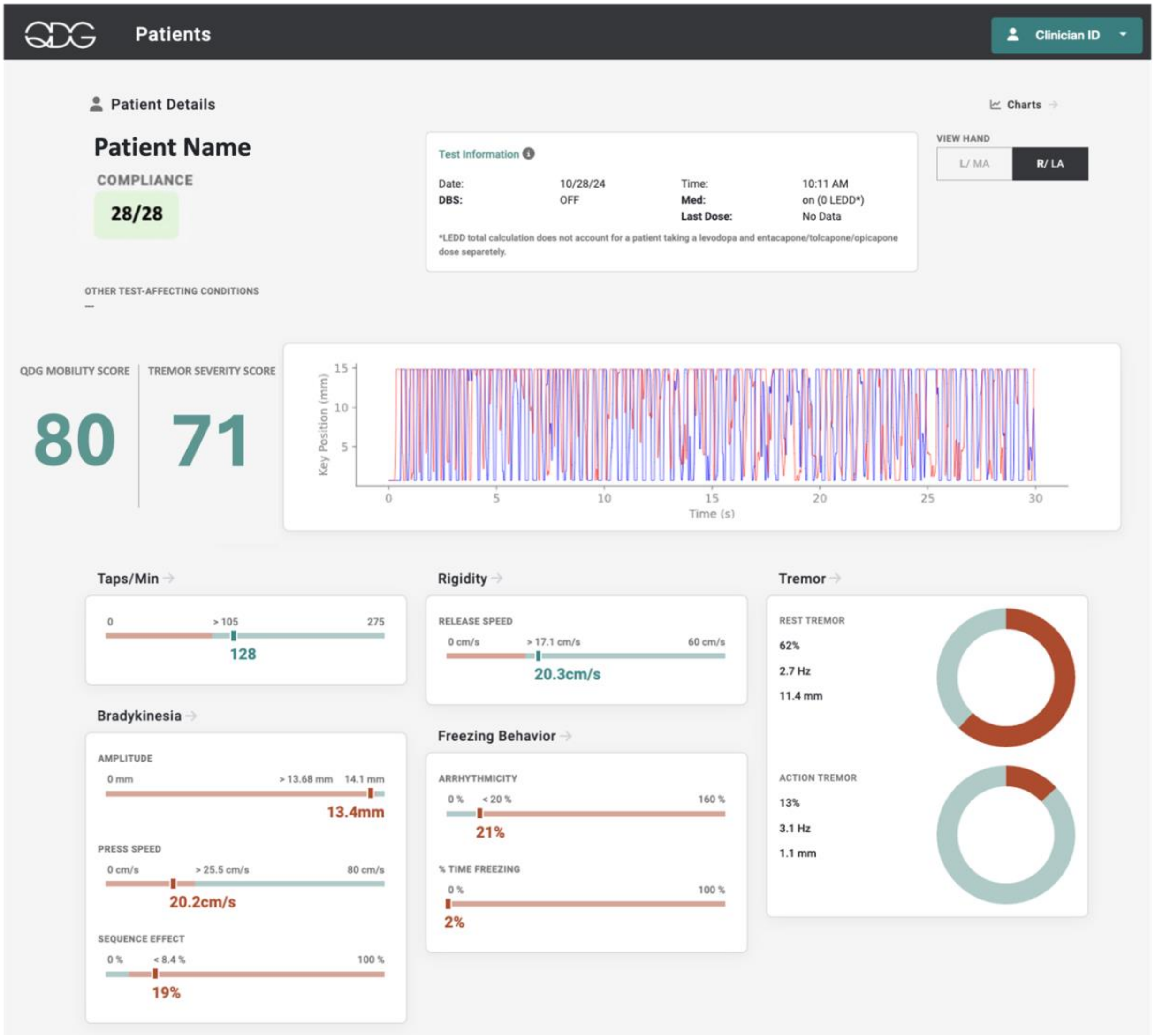


Figure S1 | QDG Clinician Dashboard View for Single Test. QDG patient information and therapy status at time of test shown above, with QDG-RAFT trace displayed for participant's more affected (MA) hand. The composite QDG Mobility Score and Tremor Severity Score are next to the trace, and the individual QDG metrics of bradykinesia, rigidity, tremor, and freezing behavior are shown below. Each metric contains a scale of values where a threshold separates the red ('abnormal') range from the blue ('normal') range. This threshold is based on the 75$^{th}$ percentile values of healthy controls in each metric.

### Experimental Protocol

*Questionnaires*

Table T1 outlines the tasks that were completed within each visit in the study protocol.

| | **In-Clinic Visit** | **At-Home Visit** | **Weekly Check-ins (1, 2, 3)** | **Exit Visit** |
|---|---|---|---|---|
| Consent | X | | | |
| MDS-UPDRS III | X | | | |
| QDG-RAFT Test | X | X | X | X |
| QDG System Training | | X | | |
| MDS-UPDRS II | | X | X | X |
| Custom Adverse Event Questionnaire | | | X | X |
| In-home Usability Testing and User Feedback Questionnaire | | | X | X |
| Design Feedback Questionnaire | | | | X |
| PD History Questionnaire | | | | X |
| Symptom Tracking and Communication Survey | | | | X |
| Exit Interview | | | | X |

Table T1 | QDG Study Task Schedule. The table delineates the tasks and questionnaires completed at each step of the study protocol through Xs.

The MDS-UPDRS II is a 13-item questionnaire, where each item is rated on a scale of 0-4 (higher number is more severe). Total MDS-UPDRS II scores were averaged across the 4 weekly check-ins for each participant.

Figure S2 displays the contents of the In-Home Usability Testing and User Feedback Questionnaire.

### QDG System In-Home Observation Visit

Goals:

1. Determine points of confusion or incorrect usage in the set-up and use of the system.
   a. Set-up, use, and breakdown of hardware.
   b. Connection of hardware and software.
   c. Navigation and use of the software application.
2. Uncover any unanticipated issues or concerns regarding use of the device.

Procedures:

1. Observe, without intervening, use from set-up to break-down. Look for unanticipated deviations from intended design.
   a. Storage location between uses:____________________________________
   b. Testing location and set-up:
      i. Location:____________________________________________
   c. Task set-up and completion:

| Set-Up | Yes | | No | |
|---|---|---|---|---|
| A. Connection of Tablet to Wifi | | | | |
| B. Hard wire connection of tablet to KeyDuo for power | | | | |
| C. Bluetooth connection of tablet to KeyDuo for data transmission | | | | |
| D. Stable seated position with KeyDuo on stable flat surface | | | | |
| E. Start new task | | | | |
| F. Navigate therapy entry screens prior to task | | | | |
| Task Performance | LEFT | | RIGHT | |
| | Yes | No | Yes | No |
| G. Used correct hand as prompted (right or left) | | | | |
| H. Device within comfortable reaching distance | | | | |
| I. Correct placement of index and middle fingers on levers | | | | |
| J. Neutral or near neutral wrist position | | | | |
| K. Used palm rest | | | | |
| L. Initiated task with repetitive alternating movement | | | | |
| M. Completed or attempted full 30 seconds of task between "go" and "stop" cues | | | | |

Comments (required for any "No" responses):________________________________________

2. Once finished, provide training and assistance as needed based on observations and answers above.
3. Discussion Guide
   a. What things work well?
   b. What things are confusing or frustrating?
   c. Did you have any problems:
      i. Connecting the tablet to the internet?
      ii. Connecting the tablet to the KeyDuo device with the cable?
      iii. Establishing or maintaining bluetooth connection between the tablet and the KeyDuo device?
4. Note any additional questions discussed or training required:________________________________________

Figure S2 | In-home usability checklist and questionnaire for QDG device testing, assessing setup, connectivity, task performance, and participant feedback on device usability and functionality.

Participants were scored on accurate execution of QDG-RAFT set-up and task performance in the In-Home Usability Testing and User Feedback Questionnaire (Figure S2). Number of observations in each sub-category of task setup and execution were totaled, and percent of correct observations was reported.

Figure S3 showcases items on the Exit Interview, which probed participants about QDG system usability.

**1. Please comment on how often you were asked to test.**

a. How easy or difficult was it to test once per day?
(0=extremely easy; 10=extremely difficult)

0 1 2 3 4 5 6 7 8 9 10

b. How easy or difficult was it to test twice per day (if applicable)?
(0=extremely easy; 10=extremely difficult)

0 1 2 3 4 5 6 7 8 9 10

Figure S3 | Exit Interview Questionnaire Items. Participants were asked to rate the ease or difficulty of testing (a) once per day and (b) twice per day, using a scale from 0 (extremely easy) to 10 (extremely difficult).

In the Exit Interview (Figure S3), participants rated their ease of use of QDG testing once and twice per day using an 11-point Likert scale (0-10, where 0=extremely easy and 10=extremely difficult). For analysis and reporting, responses were dichotomized: scores <5 were classified as 'easy' and scores >5 as 'difficult'. For visualization purposes, responses were further categorized as Extremely Easy (0-2), Moderately Easy (3-5), Moderately Difficult (6-8), or Extremely Difficult (9-10).

Figure S4 lists the contents of the Symptom Tracking and Communication Survey.

**2. How do you currently track changes in your Parkinson's disease symptoms?** Select all that apply.

☐ Write them down (e.g., calendar, paper diary, etc.)
☐ iOS or Android App (e.g., Apple Health App, APDA Symptom Tracker, Strive PD, etc.)
☐ Wearable or other remote monitoring device (e.g., Apple watch, PKG watch, etc.)
☐ Other: ______________
☐ I do not track my symptoms

**5. Please select all the ways in which you typically communicate with your physician about your Parkinson's disease symptoms.**

☐ In-person clinic visit
How often (estimate # per year)?
☐ Telehealth/video visit
How often (estimate # per year)?
☐ Phone call to physician's office
How often (estimate # per year)?
☐ Physician/provider messaging service (e.g., myHealth, MyChart, etc.)
How often (estimate # per year)?
☐ Report from wearable or other remote monitoring device
How often (estimate # per year)?
☐ Other:
How often (estimate # per year)?

6. **Has your physician previously assessed your symptoms using the Unified Parkinson's Disease Rating Scale (UPDRS)?** If so, how often do these assessments occur?

8. Remote monitoring technologies are advancing and can provide objective (i.e., numerical) measurements of movement-related symptoms in your everyday life to your physician or healthcare team. They can help track responses to different treatments or symptom progression over time.

Please rate your level of agreement with the following statement: **I wish that I had a more objective way to communicate my symptoms or changes in my symptoms to my physician/healthcare provider.**

| 0 | 1 | 2 | 3 | 4 | 5 | 6 | 7 | 8 | 9 | 10 |
|---|---|---|---|---|---|---|---|---|---|---|
| Strongly Disagree | | | | | Neutral | | | | | Strongly Agree |

Figure S4 | Symptom Tracking and Communication Survey. Survey examining Parkinson's disease patients' symptom tracking methods, healthcare provider communication channels, and attitudes toward remote monitoring technologies.

In the Symptom Tracking and Communication Survey, participants were asked whether they would prefer more objective methods of symptom communication with their provider on a scale of 0 to 10. For reporting, responses were categorized: scores <5 were classified as 'agree', scores = 5 as 'neutral', and scores >5 as 'disagree'.

### Results

### Participant Demographics

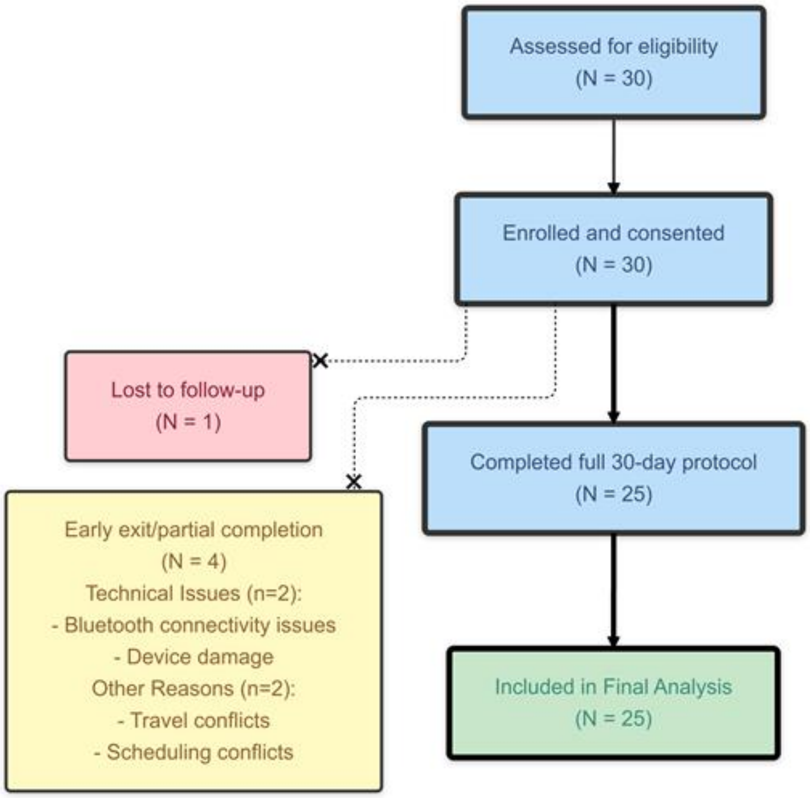


Figure S5 | Diagram of participant flow through the 30-day at-home QDG study. Of the 30 participants assessed and enrolled, 25 completed the full study duration, with 4 exiting early and 1 lost to follow-up.
